# Integrative analysis reveals regulatory effects of tandem repeat expansions in tetralogy of Fallot

**DOI:** 10.64898/2026.09.08.26362566

**Authors:** Aleksandra Mitina, Yue Yin, Robert Lesurf, Tanya Papaz, Worrawat Engchuan, Giovanna Pellecchia, Brett Trost, Thomas Nalpathamkalam, Bhooma Thiruvahindrapuram, Jade Wilson, Stephen W. Scherer, Jane Lougheed, Tapas Mondal, Mahmoud Alsalehi, Luis Altamirano-Diaz, Erwin Oechslin, Connie R. Bezzina, Alex V. Postma, David S. Winlaw, Gillian M. Blue, Seema Mital, Ryan K. C. Yuen

## Abstract

Congenital heart disease (CHD) affects approximately 1% of live births, yet a substantial proportion of cases remain genetically unexplained. Tetralogy of Fallot (TOF) is among the most common cyanotic CHDs. Tandem repeat expansions (TREs) can alter gene regulation and have been implicated in complex cardiac disease, but their contribution to structural CHD remains poorly understood.

We performed genome-wide analysis of rare TREs using short-read genome sequencing in 835 individuals with TOF and 386 controls, complemented by PacBio HiFi long-read sequencing, DNA methylation profiling, myocardial RNA sequencing, and fetal human heart single-cell RNA-seq. We further compared TRE-associated genes and biological processes between TOF and a previously characterized cardiomyopathy cohort.

We identified 1,043 rare TREs in individuals with TOF, corresponding to 1.25 rare TREs per individual, compared with 0.85 per individual in controls. TOF-associated rare TREs were enriched for GC-rich motifs, more often found in the 5′ untranslated regions (UTR; OR = 7.0, p = 2.1 × 10^−2^) and splicing regions (OR = 3.0, p = 9 × 10^−4^), and located closer to transcription start sites and splice junctions, consistent with a regulatory role. We identified 91 recurrent genic TRE loci, including recurrent 5′UTR TREs absent from controls and TREs in the CHD-associated genes *PACS1* and *TRIP4*. Long-read analysis identified 36 loci at which repeat size was significantly associated with DNA methylation, revealing both expansion-associated hypermethylation and hypomethylation. TRE-associated genes were preferentially expressed in smooth muscle cells, pericytes, and cardiomyocytes in the developing human heart. TRE-associated biological pathways were largely distinct between TOF and cardiomyopathy. Notably, rare TRE burden was higher among individuals without positive clinical genetic testing results. We estimated an excess burden of rare genic TREs of approximately 4.4% in the TOF cohort.

Rare TREs represent an underrecognized class of genetic variation contributing to TOF. Their enrichment in regulatory regions, association with locus-specific DNA methylation and transcriptional alterations, and increased burden among genetically unexplained cases support a role for TRE-mediated regulatory dysfunction in CHD pathogenesis and suggest that assessment of tandem repeat variation may improve genetic characterization of TOF.

## Introduction

Congenital heart disease (CHD) affects approximately 1% of live births and encompasses a clinically and genetically heterogeneous group of structural cardiac abnormalities. Tetralogy of Fallot (TOF) is the most common form of cyanotic CHD, accounting for 5–10% of all congenital heart defects, and results from abnormal development of the cardiac outflow tract. Although chromosomal abnormalities, copy-number variants, and rare coding variants in genes involved in cardiac development contribute to TOF, the majority of cases (>70%) remain etiologically elusive.

Tandem repeats are highly polymorphic genomic elements whose repeat number and sequence composition vary extensively among individuals. Expansion of tandem repeats can alter transcription, chromatin state, DNA methylation, RNA processing, and protein function, and pathogenic tandem repeat expansions (TREs) underlie a growing number of human disorders. Beyond established monogenic repeat expansion diseases, rare TREs have been implicated in complex disease and may act as genetic modifiers (*1–5*). We have also shown that short tandem repeat sequence composition is highly variable across human populations and is associated with gene expression, supporting a broader role for tandem repeats in gene regulation (*6*). Despite their potential regulatory effects, TREs remain poorly characterized in CHD and are not routinely assessed by conventional genetic testing.

We previously identified an increased burden of rare TREs in cardiomyopathy (CMP), including enrichment of GC-rich repeats near regulatory regions and expansion-associated DNA hypermethylation and transcriptional repression at specific loci (*5*). These findings suggested that TREs can contribute to cardiac disease through regulatory mechanisms that may not be captured by conventional coding-variant analyses. Previous genome-wide studies have further demonstrated the contribution of regulatory, copy-number, and cryptic splice variants to cardiac disease (*7*) and identified splice-disrupting variants in individuals with CHD through integration of genome sequencing with myocardial RNA sequencing (*8*). Recent work has also identified a distinct DNA methylation signature associated with pathogenic *NOTCH1* variants in CHD (*9*). Whether rare TREs similarly contribute to CHD, and whether their regulatory consequences are relevant to the developing human heart, remains unclear.

Here, we present the first genome-wide analysis of rare TREs in CHD, using a multi-site cohort of individuals with TOF and short-read genome sequencing, complemented by PacBio HiFi long-read sequencing, repeat genotyping, DNA methylation profiling, and myocardial RNA sequencing. We assessed the relationship between TRE burden and clinical characteristics and estimated the excess burden of rare genic TREs in the TOF cohort.

## Methods

### Study cohort and genome sequencing

We analyzed genome sequencing (GS) data from 835 individuals with tetralogy of Fallot (TOF) recruited across 6 Ontario sites participating in the Heart Centre Biobank Registry at the Hospital for Sick Children (Ontario, Canada), the Kids Heart BioBank at the Heart Centre for Children, The Children’s Hospital at Westmead (Sydney, Australia), and through the CONCOR registry at the Amsterdam Medical Center (Netherlands).

Institutional Research Ethics Boards of The Hospital for Sick Children, Amsterdam Medical Center, and The Children’s Hospital at Westmead gave ethical approval for the collection and use of biospecimens through the respective registries and biobanks: The Heart Centre Biobank (Ontario, Canada), CONCOR (Amsterdam, the Netherlands), and Kids Heart BioBank (Sydney, Australia). Written informed consent to participate was obtained from all participants and/or their parents or legal guardians. Study protocols adhered to the Declaration of Helsinki.

Predicted genetic ancestry was inferred using Somalier (*10*) with reference populations from the 1000 Genomes Project. Participants were assigned to European (EUR), East Asian (EAS), South Asian (SAS), African (AFR), or Admixed American (AMR) ancestry groups.

DNA was extracted from blood or saliva and prepared using Illumina TruSeq DNA PCR-Free library preparation kits, as previously described (*7*). GS was performed on Illumina HiSeq X or NovaSeq platforms, as previously described (*7*); sequencing platform information was unavailable for two participants. Sequence reads were aligned to the GRCh38/hg38 human reference genome.

For controls, we used GS data from 386 unrelated, unaffected individuals representing parents of participants recruited through the Canadian Healthy Infant Longitudinal Development (CHILD) study (*11*). The data were generated from DNA extracted from blood, prepared using a PCR-free library preparation kit, and sequenced on the Illumina HiSeq X platform using 2 × 150-bp paired-end reads. Sequence reads were aligned to the GRCh37/hg19 human reference genome, and variant coordinates were subsequently lifted over to GRCh38/hg38.

To estimate the population frequency of rare TREs, we used GS data from 2,504 participants from the 1000 Genomes Project (*12*). The data were generated from DNA derived from lymphoblastoid cell lines using PCR-free library preparation and sequenced on the Illumina NovaSeq 6000 platform with 2 × 150-bp paired-end reads. Sequence reads were aligned to the GRCh38/hg38 human reference genome.

### Detection of rare tandem repeat expansions

Tandem repeats exceeding the short-read length (150 bp) were detected from GS data using ExpansionHunter Denovo (EHdn) (*13*). EHdn output was processed using compare_anchored_irrs.py with minCount = 2. Regions were retained when at least one sample had a normalized anchored in-repeat read count (C) ≥ 2, where C = A × 40/R, A represents the raw number of anchored in-repeat reads for the region, and R represents the average sequencing depth of the sample calculated by EHdn.

Density-Based Spatial Clustering of Applications with Noise (DBSCAN) (*14*) was used to identify samples with repeat-size estimates exceeding the main cluster of observations at each region. DBSCAN was applied with a minimum cluster size (minPts) of 5 and a maximum distance between points within a cluster (epsilon) of 2. Samples identified as outliers at a given region were considered to carry a tandem repeat expansion (TRE). Rare TREs were defined as expansions occurring at a frequency <0.1% in the 1000 Genomes Project reference samples.

Within each cohort, samples for which the total number of EHdn-detected regions exceeded three standard deviations from the cohort mean were excluded from downstream analysis.

### Sequence characteristics of rare tandem repeat expansions

For each rare TRE region, the dominant repeat motif was defined as the most frequently observed motif among EHdn calls within the merged region. Motif length and GC content were calculated from the dominant motif sequence as previously described (*6*). Distributions of motif length and GC content were compared between TOF and control rare TREs using one-sided Wilcoxon rank-sum tests. The frequencies of individual repeat motifs were calculated as the proportion of rare TREs associated with each dominant motif within each cohort.

### Genomic annotation and burden analysis

Rare TRE regions were annotated according to genomic context using ANNOVAR (*15*), and TREs overlapping genic regions were assigned to the corresponding ANNOVAR-annotated gene. When a TRE overlapped multiple genomic features, annotations were prioritized in the following order: exonic, splicing (including “splicing” and “exonic; splicing”), 5′ untranslated region (5′UTR), 3′ untranslated region (3′UTR), intronic, upstream, downstream, and intergenic.

To compare the burden of rare TREs across genomic features between individuals with TOF and unaffected controls, logistic regression was performed with cohort status (TOF = 1, control = 0) as the outcome, the number of rare TREs within the genomic feature as the predictor, and the total number of rare TREs per individual as a covariate. A one-sided Wald test was used under the hypothesis of increased burden in TOF, and empirical p-values were obtained using a permutation-based approach.

Analyses were restricted to autosomal regions to avoid potential sex-chromosome bias. Analyses were performed both in the complete cohort and in a technically matched subset comprising individuals of European ancestry with DNA derived from blood and sequenced using the Illumina HiSeq X platform.

### Distance to transcription start sites

Gene coordinates and transcription start site (TSS) positions were obtained from the UCSC Genome Browser. For each TRE, the distance to the nearest TSS was calculated from the midpoint of the TRE region. Absolute distances to the nearest TSS were calculated for all detected tandem repeats and for rare TREs identified in individuals with TOF. Distributions were compared using a one-sided Wilcoxon rank-sum test under the hypothesis that rare TREs in TOF were located closer to TSSs than the background set of detected tandem repeats.

### Overlap with fetal heart epigenetic regulatory regions

Coordinates of epigenetic regulatory marks in fetal heart tissue were obtained from the Roadmap Epigenomics Consortium (*16*). Rare TRE regions were intersected with the genomic coordinates of each epigenetic mark.

To compare the burden of rare TREs overlapping each epigenetic mark between individuals with TOF and controls, two-sided Fisher’s exact tests were performed. Odds ratios and 95% confidence intervals were calculated for each epigenetic mark. Analyses were restricted to autosomal regions and were performed both in the complete cohort and in the European-ancestry subset.

### Assessment of clinical and demographic characteristics

To assess associations between clinical and demographic characteristics and rare TRE burden within the TOF cohort, four variables were examined: sex, family history of CHD, genetic testing result, and TOF subtype. Genetic testing results were classified according to the recorded clinical genetic testing interpretation as normal or abnormal. The genotype-positive group included participants with reported sequence variants, copy-number variants, or chromosomal abnormalities identified through clinical genetic testing, including findings that were not associated with, or did not fully explain, the disease phenotype. The genotype-negative group included participants whose available clinical genetic testing was reported as normal. Participants with no available genetic testing result or unknown testing status were excluded from this comparison.

For each variable, participants were divided into two groups according to the corresponding feature, and the number of rare TREs per individual was compared between groups using a two-sided Wilcoxon rank-sum test. Analyses were restricted to participants with available information for the corresponding variable; therefore, sample sizes varied among comparisons.

### Estimation of excess rare genic TRE burden

The excess burden associated with rare genic TREs to TOF was estimated by comparing the proportion of individuals carrying at least one rare genic TRE between TOF cases and controls. To account for technical differences in TRE detection between cohorts, the proportion of TOF cases carrying a rare genic TRE was adjusted using the relative occurrence of intergenic TREs in TOF cases and controls. The estimated excess burden associated with rare genic TREs was calculated as the difference between the adjusted proportion of TOF cases carrying rare genic TREs and the observed proportion of controls carrying rare genic TREs.

### Recurrent TREs and disease-gene enrichment

Recurrent TRE-associated genes were defined as genes harbouring rare TREs in two or more individuals with TOF. Recurrent TRE-associated genes were compared with a previously curated set of Tier 1 CHD genes with definitive evidence for association with CHD based on human genetic and clinical evidence (*17*), and with genes implicated in established repeat expansion disorders.

Enrichment within disease-gene sets was assessed using one-sided Fisher’s exact tests, with all genes in the genome used as the background. Odds ratios and corresponding p-values were calculated for each comparison.

Recurrent non-intronic TRE loci were ranked according to their frequency in TOF and controls.

### Gene interaction and functional enrichment analysis

Genes associated with rare TREs in individuals with TOF were analyzed using the GeneMANIA Cytoscape app (*18*). Network construction was restricted to physical and pathway interactions, with network weighting based on Gene Ontology (GO) Biological Process annotations. Genes with GO annotations and at least one interaction represented in the network database were used as the background gene set.

GeneMANIA functional enrichment analysis was restricted to GO terms containing between 10 and 300 annotations and excluded annotations inferred from reviewed computational analysis (RCA) or electronic annotation (IEA) (*19*). Multiple-testing correction was performed using the Benjamini–Hochberg procedure, and terms with a false discovery rate (FDR) q-value ≤0.1 were retained.

### Fetal human heart single-cell RNA-seq data

Publicly available fetal human heart single-cell RNA sequencing data from Knight-Schrijver et al. (2022; GSE216019) were used (*20*). The dataset comprised 30,872 fetal cardiac cells from seven donors collected at 8–12 weeks post-conception and was obtained as a pre-processed and annotated Seurat object.

To characterize the cellular expression of genes harbouring rare TREs in the TOF cohort, expression scores for TRE-associated genes were calculated for individual cells using the AddModuleScore function in Seurat. Scores represented the mean expression of TRE-associated genes relative to randomly selected expression-level-matched control genes. Scores were calculated for the full TOF TRE-associated gene set (n = 433), restricted to genes represented in the fetal heart single-cell RNA-seq dataset.

### Long-read genome sequencing

PacBio HiFi long-read genome sequencing was performed in a subset of 43 individuals with TOF, as previously described (*21*). Briefly, high-molecular-weight DNA was used for library preparation and sequencing on the PacBio Sequel IIe platform using 8M SMRT Cells at The Centre for Applied Genomics (TCAG), The Hospital for Sick Children, Toronto, Canada.

Unaligned BAM files were processed using the PacBio Human GS Workflow, and reads were aligned to the GRCh38 human reference genome.

### Selection of tandem repeat loci and TRGT catalogue matching

TOF tandem repeat regions identified using EHdn were intersected with tandem repeat loci from the genome-wide Adotto TRGT catalogue (*22*). Each TOF region was assigned a dominant motif representing the most frequently observed motif among EHdn calls within the merged region.

Repeat motifs were normalized for cyclic rotations and reverse-complement sequences before comparison with motifs from overlapping TRGT catalogue entries. Closely related non-identical motifs were additionally compared by sequence similarity to identify catalogue loci representing the same or a highly similar repeat sequence. For TOF regions overlapping multiple catalogue entries, candidate matches were evaluated according to motif concordance and genomic overlap. Selected TRGT loci were used to construct a custom catalogue for long-read tandem repeat analysis.

Original TOF-region coordinates, genomic annotations, associated genes, dominant motifs, and alternative motifs identified by EHdn were retained for downstream analyses.

### TRGT repeat-size and DNA methylation analysis

Selected tandem repeat loci were genotyped from PacBio HiFi sequencing data using Tandem Repeat Genotyping Tool (TRGT) (*23*). Allele-specific repeat-size, repeat-purity, and DNA methylation estimates were obtained for each locus and individual. Repeat purity represents the proportion of bases within the repeat allele that match the repeat motif, with higher values indicating a more homogeneous repeat sequence. Analyses were restricted to samples with available measurements for the variables being compared.

For each sample and locus, repeat size was calculated as the sum of the sizes of the two alleles, while repeat purity and methylation were calculated as the mean values across the two alleles. The association between repeat size and DNA methylation or repeat purity was assessed across individuals using Pearson correlation. For each locus, the Pearson correlation coefficient, nominal p-value, and number of samples with available data were recorded. Nominal statistical significance was defined as p ≤ 0.05.

### RNA sequencing and gene-expression analysis

RNA-seq data from 159 TOF individuals with available myocardial tissue were generated and processed as previously described (*8*). Briefly, RNA-seq was performed on myocardial tissue obtained during cardiac surgery or transplantation and sequenced using the Illumina HiSeq 2500 platform at The Centre for Applied Genomics, The Hospital for Sick Children, Toronto, Canada. Gene expression was quantified as transcripts per million (TPM).

To assess gene-expression differences associated with methylation-linked TREs, loci showing a significant association between repeat size and DNA methylation in the TRGT analysis were mapped to their corresponding EHdn regions and associated genes. For loci with RNA-seq data available for the corresponding gene, individuals were classified as EHdn-positive or EHdn-negative according to the presence or absence of an EHdn-detected expansion at that locus. TPM values were compared between EHdn-positive and EHdn-negative individuals using a two-sided Wilcoxon rank-sum test.

Among EHdn-positive individuals with both repeat-size estimates and RNA-seq data, associations between EHdn-estimated repeat size and gene expression were assessed using Pearson correlation. The Pearson correlation coefficient, p-value, and number of samples were recorded for each locus.

### Locus-specific repeat genotyping with ExpansionHunter

For selected candidate regions identified by EHdn, tandem repeat coordinates were refined using the Simple Repeat track from the UCSC Genome Browser (*24,25*). Refined repeat coordinates and corresponding repeat motifs were used to construct a custom ExpansionHunter variant catalogue. ExpansionHunter was applied to short-read GS data to obtain locus-specific repeat-size estimates for both alleles across individuals.

### Comparison of TRE-associated genes and pathways between TOF and cardiomyopathy

TRE-associated genes identified in the TOF cohort (n=433) were compared with TRE-associated genes previously identified in the cardiomyopathy cohort (n=127) (*5*). Genes were classified as unique to TOF, unique to cardiomyopathy, or shared between the two cohorts.

Functional interaction networks and enriched biological processes for the TOF-associated gene set were generated as described above. Cardiomyopathy-associated functional networks and enriched biological processes were obtained from the previously described analysis (*5*). Shared TRE-associated genes represented in enriched biological processes in both disease cohorts were identified.

## Results

### Rare tandem repeat expansions are increased in individuals with TOF

We identified 1,043 rare tandem repeat expansions (TREs), defined as expansions occurring at a frequency <0.1% in the 1000 Genomes Project reference samples, across 835 individuals with TOF, corresponding to an average of 1.25 rare TREs per individual (***Supplementary Table 1***). In comparison, 330 rare TREs were identified across 386 controls, corresponding to an average of 0.85 rare TREs per individual. Rare genic TREs were identified in 347 of 835 individuals with TOF (41.6%) and 127 of 386 controls (32.9%; OR = 1.45, two-sided Fisher’s exact test, p = 4.45 × 10^−3^). Intergenic TREs were detected in 386 of 835 individuals with TOF (46.2%) and 160 of 386 controls (41.5%). After correction for differences in background intergenic TRE rates between the cohorts, the estimated excess burden associated with rare genic TREs was 4.4% in the TOF cohort.

### Rare TREs in TOF are enriched for GC-rich repeat motifs

We next examined the sequence characteristics of rare TREs identified in individuals with TOF and controls (**Figure 1**). The distribution of dominant repeat motif lengths was similar between the two cohorts, with no significant difference in motif length (one-sided Wilcoxon rank-sum test, p = 0.92; **Figure 1a**). The corresponding motif-length frequency distribution in controls is shown in ***Supplementary Figure 1a***.

Rare TREs identified in TOF had significantly higher motif GC content than those identified in controls (one-sided Wilcoxon rank-sum test, p = 3.5 × 10^−8^; **Figure 1b**). The frequency distributions showed a greater representation of GC-rich TREs in TOF, whereas controls showed a comparatively greater representation of lower-GC-content TREs (***Supplementary Figure 1b***). Among the most frequently observed repeat motifs were AAAG, AAG, AC, AAAAT, and AT (**Figure 1c**). Comparison of individual motif frequencies identified three motifs that differed significantly between TOF and controls after multiple-testing correction: AC was enriched in TOF rare TREs (8.17% vs. 0.72%; OR = 12.3, FDR = 2.9 × 10^−8^), whereas AT (5.01% vs. 18.7%; OR = 0.23, FDR = 3.7 × 10^−14^) and AATGG (1.58% vs. 9.57%; OR = 0.15, FDR = 1.5 × 10^−10^) occurred at higher frequencies among control rare TREs.

**Figure 1.**
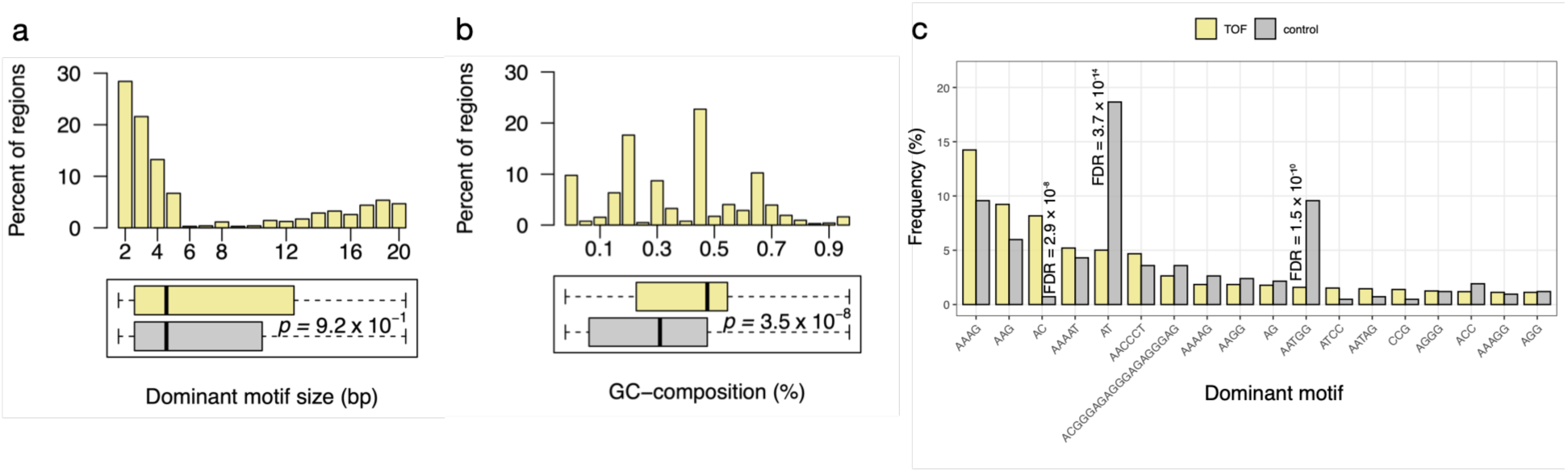
Sequence characteristics of rare tandem repeat expansions in tetralogy of Fallot. **(a)** Distribution of dominant repeat motif lengths among rare tandem repeat expansions (TREs) identified in individuals with tetralogy of Fallot (TOF; yellow) and controls (grey). Histograms show the percentage of rare TREs in each motif-length category; boxplots summarize the motif-length distributions. The p-value is from a one-sided Wilcoxon rank-sum test. **(b)** Distribution of dominant motif GC content among rare TREs in TOF (yellow) and controls (grey). Histograms show the percentage of rare TREs in each GC-content bin; boxplots summarize the GC-content distributions. The p-value is from a one-sided Wilcoxon rank-sum test. **(c)** Frequencies of the most common dominant repeat motifs represented in both TOF and control cohorts, shown as the percentage of rare TREs within each cohort. FDR values are from two-sided Fisher’s exact tests with Benjamini-Hochberg correction across all dominant motifs tested. Motifs are ordered by frequency in the TOF cohort.

### Rare TREs in TOF are enriched in 5′UTRs and splicing regions and occur closer to transcription start sites

We next examined the genomic distribution of rare TREs in the TOF cohort (Figure 2). Compared with controls, rare TREs in TOF were significantly enriched in 5′ untranslated regions (UTRs; OR = 7.0, p = 2.1 × 10^−2^) and splicing regions (OR = 3.0, p = 9 × 10^−4^; **Figure 2a**; ***Supplementary Table 2***). To account for potential technical and ancestry-related differences between the TOF and control cohorts, we repeated the analysis in a subset restricted to individuals of European ancestry with blood-derived DNA sequenced on the Illumina HiSeq X platform, and the same enrichment pattern was observed (***Supplementary Figure 2; Supplementary Table 3***).

Rare TREs in TOF were located significantly closer to transcription start sites (TSSs) than the background set of detected tandem repeats (one-sided Wilcoxon rank-sum test, p = 3.9 × 10^−2^; **Figure 2b**) and significantly closer to splice junctions than rare TREs in controls (one-sided Wilcoxon rank-sum test, p = 2.5 × 10^−2^; **Figure 2c**).

### Rare TRE burden across fetal heart epigenetic regulatory regions

We assessed the burden of rare TREs overlapping fetal heart epigenetic regulatory marks obtained from the Roadmap Epigenomics Consortium (*16*) in TOF compared with controls. In the complete cohort, including individuals of all ancestries, rare TREs were significantly enriched in DNase I hypersensitive sites identified by MACS2 (OR = 1.5, p = 1.5 × 10^−2^) and H3K4me1 active enhancer regions (OR = 1.5, p = 2.7 × 10^−2^). In the European-ancestry subset, significant enrichment was observed for DNase I hypersensitive sites (OR = 1.6, p = 7.1 × 10⁻^3^), and H3K36me3 actively transcribed regions (OR = 1.6, p = 4.3 × 10^−2^; ***Supplementary Figures 3–4***; ***Supplementary Tables 4–5***).

### TRE-associated genes are enriched in developmental and cellular signaling pathways

To examine the biological processes represented among genes overlapping rare TREs in TOF, we performed interaction-network and functional enrichment analyses (**Figure 2d**). Enriched biological processes included axonogenesis, developmental growth, calcium ion transport, and regulation of transporter activity.

Cardiac-related genes represented within these enriched processes included the TRE-associated genes *SEMA3C*, *SLIT3*, *ROBO2*, *CACNB1*, *CACNB2*, *TRPC6*, *PRKCA*, *PLCB1*, *PAK1*, *MYLK*, *IGF1*, *CAV1*, *NTN1*, and *SEMA3G*, as well as *RYR2* and *PDE4D*, which directly interact with these genes (**Figure 2d**). TRE-associated genes showed a trend toward enrichment among Tier 1 CHD genes (*17*) (OR = 1.50, one-sided Fisher’s exact test, p = 8.2 × 10^−2^).

**Figure 2.**
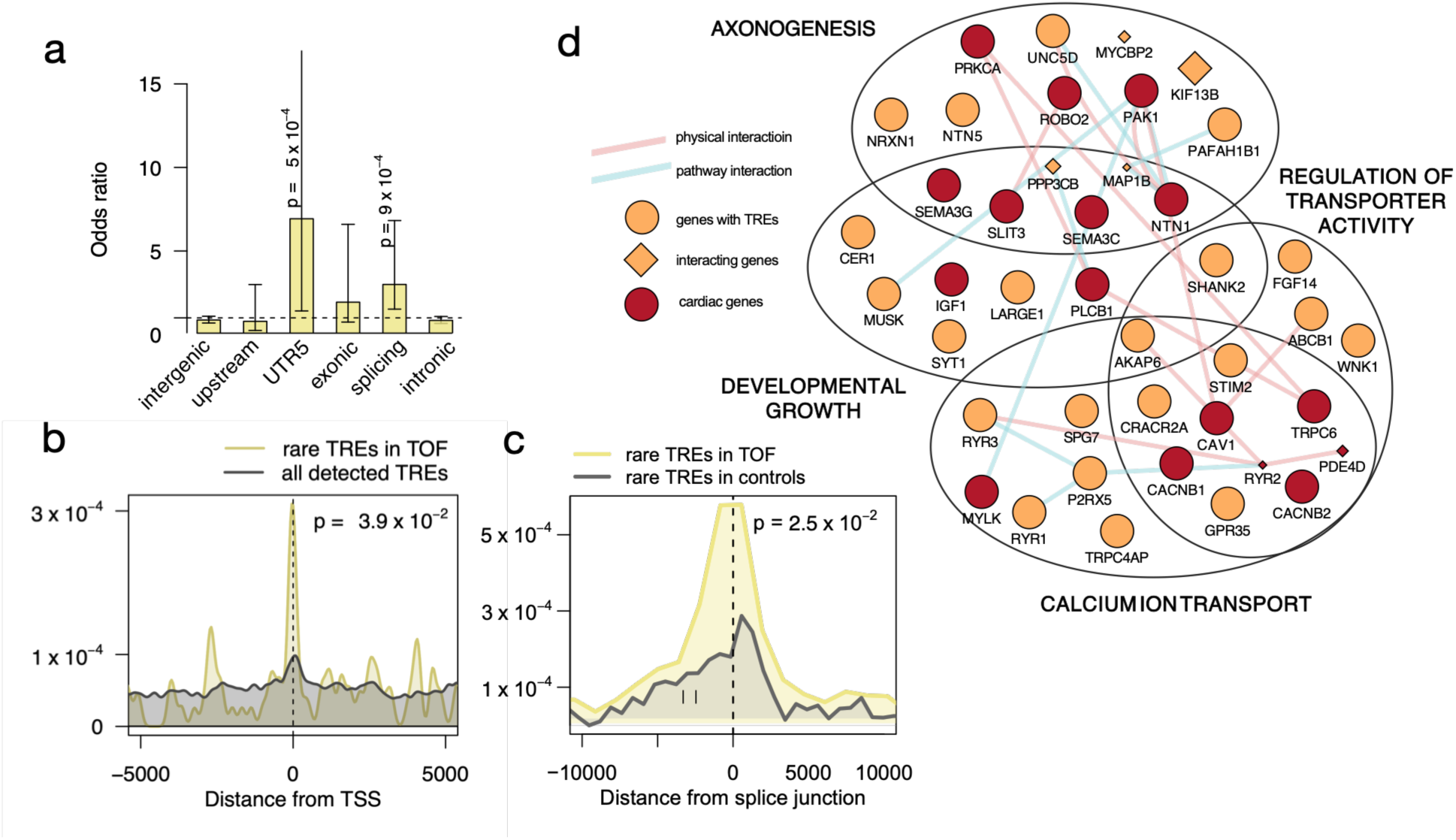
Genomic distribution and functional characteristics of rare tandem repeat expansions in tetralogy of Fallot. **(a)** Burden of rare TREs across genomic features in individuals with TOF compared with controls. Bars represent odds ratios from logistic regression adjusted for the total number of rare TREs per individual; error bars indicate 95% confidence intervals. The horizontal dashed line indicates an odds ratio of 1. P-values are shown for significant comparisons. **(b)** Distribution of absolute distances to the nearest transcription start site (TSS) for all detected tandem repeats (grey) and rare TREs identified in individuals with TOF (yellow). The p-value is from a one-sided Wilcoxon rank-sum test. **(c)** Distribution of distances to the nearest splice junction for rare TREs identified in controls (grey) and individuals with TOF (yellow). Negative and positive distances indicate distances to acceptor and donor splice sites, respectively. The vertical dashed line indicates the splice junction. The p-value is from a one-sided Wilcoxon rank-sum test of absolute distances. **(d)** Interaction network of genes associated with rare TREs and enriched biological processes. Circles represent TRE-associated genes and diamonds represent directly interacting genes. Dark red indicates cardiac-related genes, and orange indicates other genes. Edges indicate physical interactions (pink) or pathway relationships (blue).

### Recurrent TREs identify candidate loci in TOF

We next examined TRE loci recurrently observed in the TOF cohort. In total, 91 recurrent genic TRE loci were identified in individuals with TOF (***Supplementary Table 6***). Among recurrent genic TREs, five 5′UTR loci were observed in more than one individual with TOF and were absent from controls, including those in *BCL2L11*, *FAM149A*, *TBC1D7-LOC100130357*, *ZNF713*, and *LRRC6* (**Table 1**). Locus-specific repeat-size distributions estimated using ExpansionHunter are shown in ***Supplementary Figure 5***.

**Table 1.** Recurrent 5′UTR tandem repeat loci identified in individuals with TOF.

| Coordinate and dominant motif | TOF (n) | Controls (n) | Gene (region) | OMIM phenotype |
| --- | --- | --- | --- | --- |
| chr2:111120569–111121320 (CCG) | 2 | 0 | *BCL2L11* | - |
| chr4:186144435–186145322 (CCCGCG) | 2 | 0 | *FAM149A* | - |
| chr6:13328020–13328949 (AGCGGC) | 2 | 0 | *TBC1D7-LOC100130357* | TBC1D7 deficiency syndrome* |
| chr7:55887151–55888103 (CCG) | 2 | 0 | *ZNF713* | - |
| chr8:132673484–132674654 (AGG) | 2 | 0 | *LRRC6* | Primary ciliary dyskinesia |

Among the complete set of recurrent genic TREs, two genes, *PACS1* and *TRIP4*, overlapped the curated Tier 1 CHD gene set (*17*). An intronic TRE in *PACS1* was observed in four individuals with TOF and no controls, and an intronic TRE in *TRIP4* was observed in three individuals with TOF and no controls (***Supplementary Table 6***).

### Tandem repeat size is associated with DNA methylation and gene expression

We next examined the relationship between large TREs and gene expression using available myocardial RNA-seq data from 159 individuals with TOF. Among 294 TRE–gene pairs with sufficient RNA-seq data for comparison between individuals with and without a large repeat, 23 showed significant differences in gene expression after Benjamini–Hochberg correction (FDR < 0.05), representing 22 unique genes. Of these, 22 (95.7%) showed lower expression in individuals with a large repeat, whereas one (4.3%) showed higher expression. Genes showing reduced expression included *P2RX5* (p = 1.68 × 10^−4^), *STAT3* (p = 2.05 × 10^−4^), *CRIP2* (p = 3.84 × 10^−4^), *GRK5* (p = 1.05 × 10^−3^), *PRKCA* (p = 1.13 × 10^−3^), and *CDON* (p = 1.45 × 10^−3^; Supplementary Figure 6; Supplementary Table 7).

To assess the relationship between tandem repeat size and DNA methylation, we analyzed TRGT-derived repeat size and methylation measurements from PacBio HiFi sequencing data in a subset of 43 individuals with TOF (***Supplementary Table 8***). Among 555 fine-mapped loci with both repeat size and DNA methylation measurements available, 36 showed a significant correlation between repeat size and DNA methylation (**Figure 3a**; ***Supplementary Table 9***).

Significant correlations were observed in both directions, with increasing repeat size associated with increased methylation at 18 loci and decreased methylation at 18 loci.

At the *P2RX5* locus, increasing repeat size was associated with increased DNA methylation (Pearson r = 0.34, p = 4.24 × 10^−2^, n = 36; **Figure 3b**). Increasing repeat size was also associated with greater repeat sequence purity, reflecting a higher proportion of the repeat allele matching the repeat motif (Pearson r = 0.29, p = 9.12 × 10^−2^, n = 36; **Figure 3c**). Individuals with a large repeat had significantly lower *P2RX5* expression than individuals without a large repeat (Wilcoxon rank-sum test, p = 1.7 × 10⁻^4^; **Figure 3d**).

**Figure 3.**
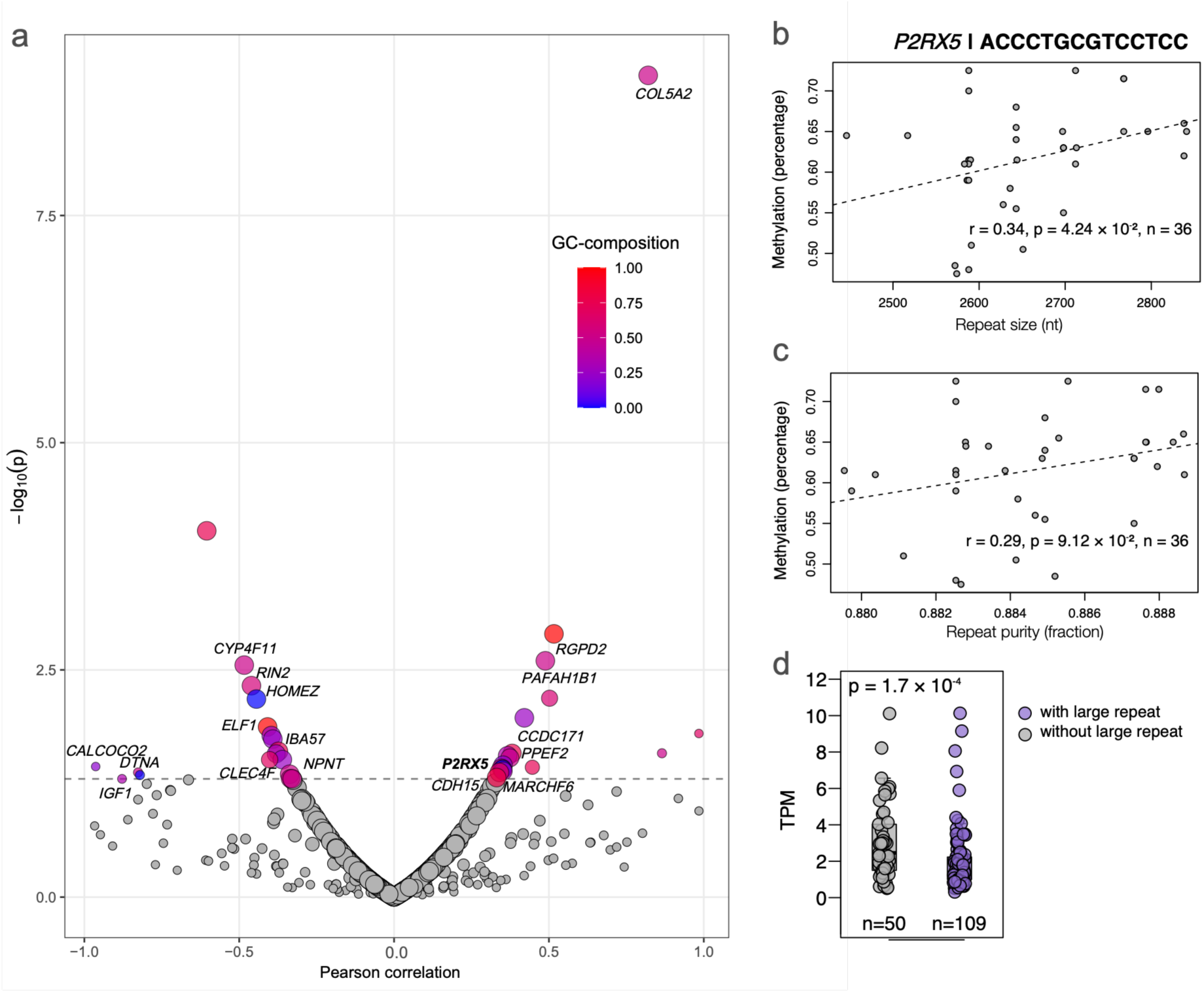
Tandem repeat size, DNA methylation, and gene expression. **(a)** Associations between tandem repeat size and DNA methylation measured from PacBio HiFi sequencing data. Each point represents a tandem repeat locus; the x-axis shows the Pearson correlation coefficient between TRGT-derived repeat size and DNA methylation, and the y-axis shows −log10(p-value). Points are coloured according to repeat GC content. The horizontal dashed line indicates p = 0.05. **(b)** Relationship between TRGT-derived repeat size (x-axis) and DNA methylation (y-axis) at the *P2RX5* locus. **(c)** Relationship between TRGT-derived repeat size (x-axis) and DNA purity (y-axis) at the *P2RX5* locus. **(d)** *P2RX5* expression, measured as transcripts per million (TPM), in individuals with (purple) and without (grey) a large repeat, as identified by EHdn. The p-value is from a two-sided Wilcoxon rank-sum test. In **(b, c)**, dashed lines indicate linear regression fits; Pearson correlation coefficients (r), p-values, and sample sizes (n) are shown. EHdn, ExpansionHunter Denovo; TRGT, Tandem Repeat Genotyping Tool.

### TRE-associated genes show cell type-specific expression in the developing human heart

To examine the cellular expression of TRE-associated genes during heart development, we analyzed fetal human heart single-cell RNA-seq data comprising 30,872 cells from seven donors without CHD, collected at 8–12 weeks post-conception (*20*). TRE-associated genes showed the highest expression relative to expression-matched control genes in smooth muscle cells, pericytes, and cardiomyocytes (**Figure 4a**; ***Supplementary Table 10***). Visualization of TRE-associated gene expression scores across individual cells showed their distribution among the major fetal cardiac cell populations (**Figure 4b**).

**Figure 4.**
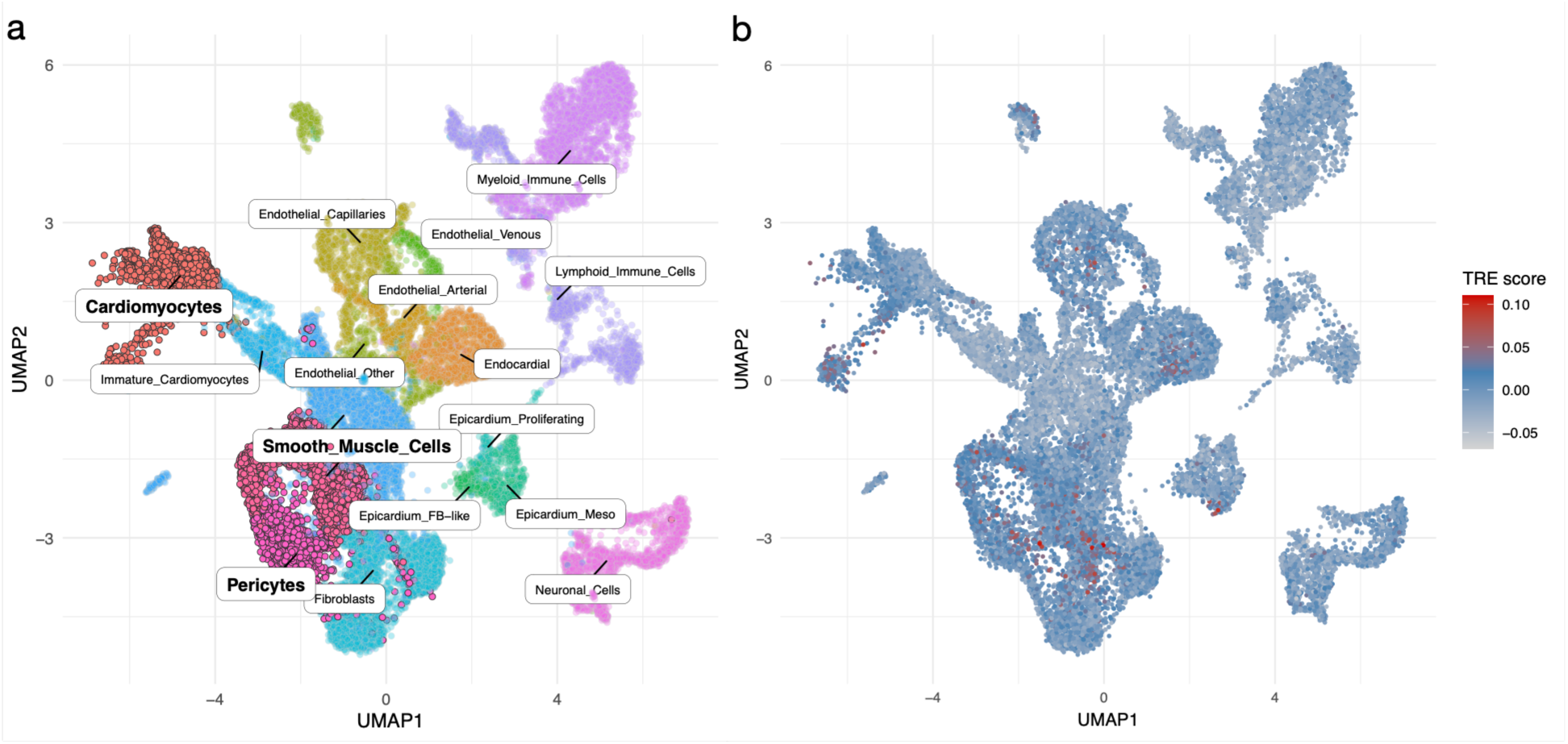
Expression of TOF TRE-associated genes across fetal human cardiac cell types. **(a)** Cell types with the highest mean TRE gene expression scores, with smooth muscle cells, pericytes, and cardiomyocytes highlighted. **(b)** Single-cell TRE gene expression scores representing the mean expression of 433 TRE-harbouring genes relative to expression-matched background genes, computed using Seurat’s AddModuleScore. The colour scale ranges from low (grey) to high (red). Fetal human heart single-cell RNA-seq data were obtained from Knight-Schrijver et al. (2022; GSE216019).

### TRE-associated genes and biological pathways in TOF and cardiomyopathy

To assess the relevance of rare TREs to TOF and determine whether they overlap with TREs implicated in other cardiac conditions, we compared the 433 TRE-associated genes identified in TOF with 127 TRE-associated genes previously identified in cardiomyopathy (5). Forty-five genes were shared between the two cohorts, whereas 388 were unique to TOF and 82 were unique to cardiomyopathy (***Supplementary Figure 7a***; ***Supplementary Table 11***).

We next compared the enriched biological processes associated with TRE genes in the two cohorts (***Supplementary Figure 7b***). The enriched processes did not overlap between TOF and cardiomyopathy. Among TRE-associated genes represented within these processes, only two genes, *ROBO2* and *SEMA3C*, were shared between the cohorts. *ROBO2* was represented in axonogenesis in TOF and in heart growth and heart valve morphogenesis in cardiomyopathy, whereas *SEMA3C* was represented in axonogenesis and developmental growth in TOF and in heart growth in cardiomyopathy (***Supplementary Figure 7b***).

### Rare TRE burden across clinical and demographic characteristics

We assessed whether rare TRE burden differed according to clinical and demographic characteristics within the TOF cohort. Individuals with a genotype-negative clinical genetic testing result had a significantly higher rare TRE burden than those with a genotype-positive result (p = 1.8 × 10^−2^; **Figure 5**). Rare TRE burden did not differ significantly by sex, family history, or TOF subtype (two-sided Wilcoxon rank-sum tests; ***Supplementary Table 12***).

**Figure 5.**
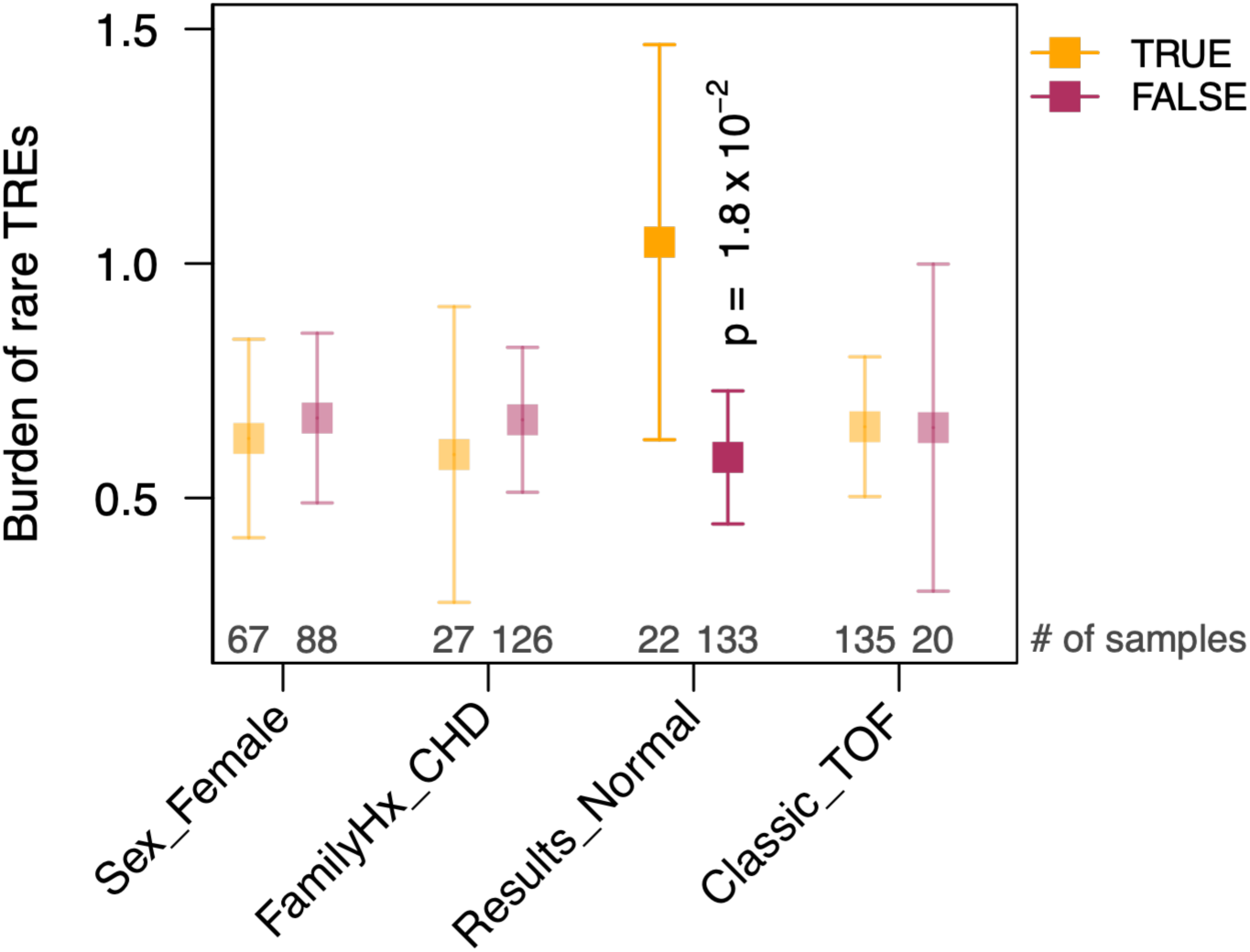
Clinical features of rare tandem repeat expansions. Comparison of the number of rare TREs (y-axis, 95% confidence interval) with respect to four variables (x-axis) in individuals with (orange) or without (magenta) the indicated feature. Two-sided Wilcoxon’s rank sum test p-value is labelled for the variables with significant difference between the two groups.

## Discussion

In this study, we investigated the contribution of rare TREs to the genetic architecture of TOF using an integrative approach combining short- and long-read genome sequencing, DNA methylation, transcriptomic, and fetal heart single-cell RNA-sequencing data. We found an increased burden of rare TREs in individuals with TOF compared with unaffected controls, with preferential localization to 5′ untranslated and splicing regions and closer proximity to transcription start sites and splice junctions, suggesting that their contribution may be mediated in part through effects on gene regulation. TRE-associated genes converged on developmental and cardiac-relevant biological processes and were preferentially expressed in smooth muscle cells, pericytes, and cardiomyocytes during early fetal heart development. Integration of long-read methylation profiling with myocardial RNA-seq data further identified repeat-associated changes in both DNA methylation and gene expression, demonstrating multiple potential regulatory consequences of repeat variation. Together, these findings expand the spectrum of genetic variation potentially contributing to TOF and support rare TREs as a class of regulatory variation relevant to CHD.

Several of the recurrent loci were GC-rich repeats and located in 5′UTRs, including *BCL2L11*, *FAM149A*, *TBC1D7-LOC100130357*, and *ZNF713*. Notably, *LRRC6* is a motile-cilia gene implicated in laterality defects and congenital heart abnormalities (*26, 27*). The broader set of recurrent TRE-associated genes also included *PACS1* and *TRIP4*, both established CHD-associated genes. *PACS1* is involved in intracellular protein trafficking and is associated with a multisystem developmental disorder that can include CHD (*28, 29*), while biallelic loss-of-function variants in *TRIP4* cause severe congenital neuromuscular disorders in which cardiac abnormalities, including septal defects and cardiomyopathy, have been reported (*30–32*). The recurrence of TREs in established disease genes, together with recurrent 5′UTR CGG expansions in additional loci, identifies specific candidates for further investigation of repeat-mediated effects on gene regulation and TOF risk.

Long-read sequencing and RNA-seq provided evidence that a subset of the identified TREs may have functional consequences. Gene expression was significantly reduced in individuals with large repeats at 22 of 23 TRE–gene pairs showing significant expression differences, suggesting that reduced expression is a frequent transcriptional feature associated with large TREs in this dataset. At the *P2RX5* locus, increasing repeat size was associated with increased DNA methylation, while individuals with a large repeat showed reduced *P2RX5* expression, consistent with an expansion-associated hypermethylation and transcriptional repression mechanism.

*P2RX5* is expressed in cardiac tissue, with P2X5 receptor expression previously demonstrated in both atrial and ventricular myocardium and particularly prominent during early postnatal heart development (*33*). More broadly, repeat size was associated with both increased and decreased methylation across the fine-mapped TRE loci, indicating that the epigenetic consequences of repeat expansion may vary by locus.

Comparison with our previous cardiomyopathy cohort revealed overlap at the gene level, with 45 TRE-associated genes shared between TOF and cardiomyopathy. However, the enriched biological processes identified in the two cohorts did not overlap, suggesting that TREs may contribute to TOF and cardiomyopathy through distinct biological mechanisms. Consistent with this, among the genes contributing to the enriched biological processes in each cohort, only two genes, *ROBO2* and *SEMA3C*, were shared. Both genes have established roles in cardiac development, with *ROBO2* implicated in cardiac septation and *SEMA3C* required for cardiac outflow tract development and septation (*34, 35*).

Finally, after accounting for differences in background intergenic TRE detection between cases and controls, we estimated an excess burden associated with rare genic TREs of approximately 4.4% in the TOF cohort. Rare TRE burden was also significantly higher in individuals with normal clinical genetic testing results, whereas no differences were observed by sex, family history, or TOF subtype. This finding further suggests that rare TREs may represent an additional source of genetic variation in TOF, particularly among individuals without findings from conventional clinical genetic testing.

Several limitations should be considered. TRE detection from short-read sequencing remains challenging, particularly for large expansions and repetitive regions, and differences in ancestry, DNA source, and sequencing platform between cases and controls may introduce technical variability despite our sensitivity analyses. In addition, functional consequences could be assessed only for subsets of TREs with available long-read sequencing, DNA methylation, RNA-seq, or relevant developmental expression data. Together, these findings support rare TREs as a previously underrecognized contributor to the genetic architecture of TOF and demonstrate their potential regulatory effects on genes relevant to cardiac development.

## Data Availability

Data generated and analyzed in this study are available from the corresponding authors upon reasonable request.

## Declarations

### Data and code availability

Data generated and analyzed in this study and analysis code are available from the corresponding authors upon reasonable request.

### Ethics approval and consent to participate

Institutional Research Ethics Boards of The Hospital for Sick Children, Amsterdam Medical Center, and The Children’s Hospital at Westmead approved the collection and use of biospecimens through the respective registries and biobanks: the Heart Centre Biobank (Ontario, Canada), CONCOR (Amsterdam, the Netherlands), and Kids Heart BioBank (Sydney, Australia). Written informed consent to participate was obtained from all participants and/or their parents or legal guardians. All study protocols adhered to the Declaration of Helsinki.

### Author contributions

Conceptualization: SM, RKCY.

Methodology: WE, BTrost, RL, BThiruvahindrapuram, TN, RKCY.

Investigation: AM, YY, GP.

Visualization: AM, YY, WE, GP.

Data curation: SWS, JL, TM, MA, LAD, EO, AVP, GMB, DSW, CRB.

Funding acquisition: SM, RKCY.

Project administration: TP, JW.

Supervision: SM, RKCY.

Writing—original draft: AM, RKCY.

Writing—review & editing: SM, WE, RL, TP, AVP.

RKCY and AM accessed and verified the underlying data. All authors read and approved the final version of the manuscript.

