## Supplementary Figures for "Integrative analysis reveals regulatory effects of tandem repeat expansions in tetralogy of Fallot"

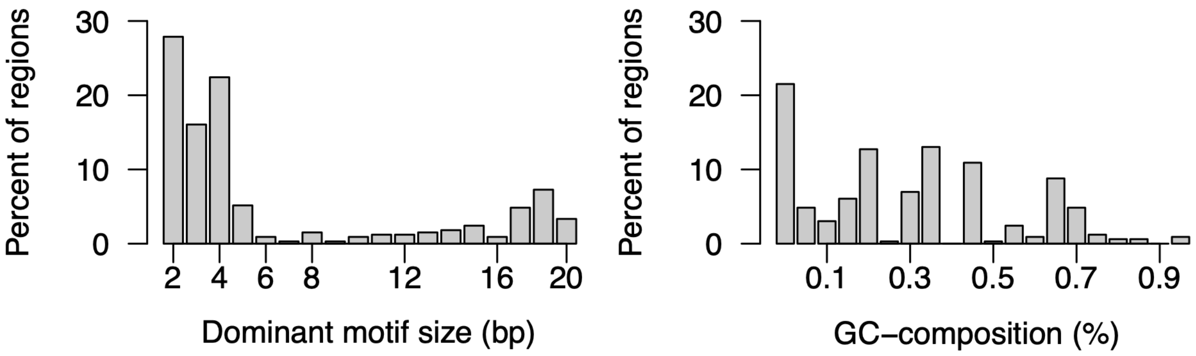


**Supplementary Figure 1. Sequence characteristics of rare tandem repeat expansions in controls.** (a) Frequency distribution of dominant repeat motif lengths among rare tandem repeat expansions (TREs) identified in controls. Bars represent the percentage of control rare TREs within each motif-length category. (b) Frequency distribution of dominant motif GC content among rare TREs identified in controls. Bars represent the percentage of control rare TREs within each GC-content bin.

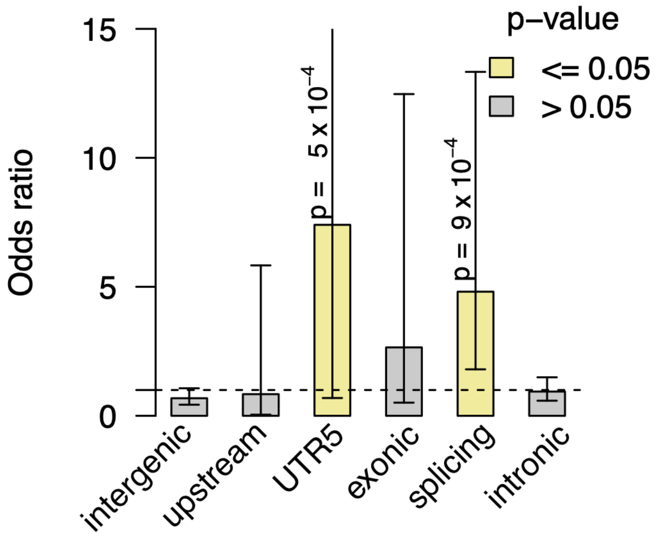


**Supplementary Figure 2. Rare TRE burden across genic regions in a technically and ancestrally matched TOF subset.** Burden of rare TREs across genic regions in individuals with TOF of European ancestry whose DNA was derived from blood and sequenced on the Illumina HiSeq X platform, compared with controls. Bars represent odds ratios from logistic regression adjusted for the total number of rare TREs per individual; error bars indicate 95% confidence intervals. The horizontal dashed line indicates an odds ratio of 1. P-values are shown for significant comparisons.

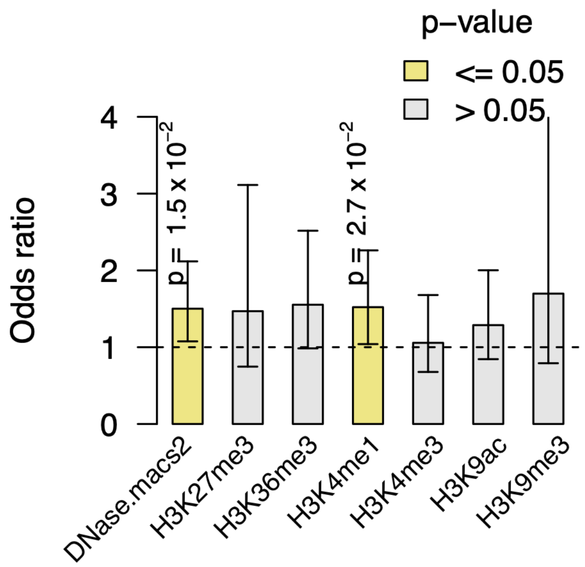


**Supplementary Figure 3. Rare TRE burden across fetal heart epigenetic regulatory regions.** Burden of rare TREs overlapping fetal heart epigenetic marks in individuals with TOF compared with controls. Bars represent odds ratios from Fisher's exact tests; error bars indicate 95% confidence intervals. The horizontal dashed line indicates an odds ratio of 1.

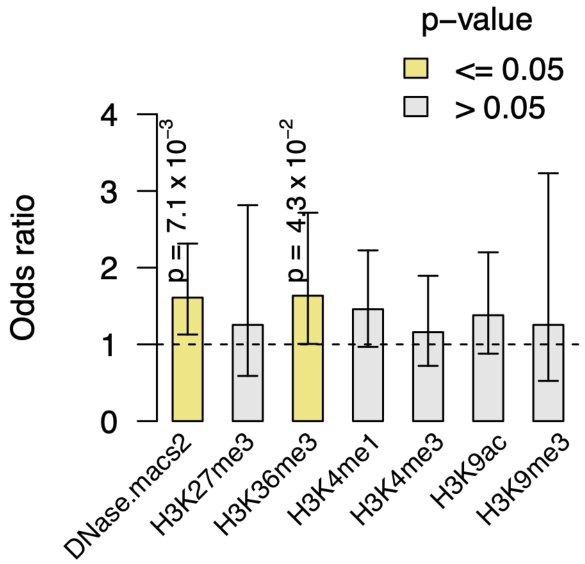


**Supplementary Figure 4. Rare TRE burden across fetal heart epigenetic regulatory regions in the European-ancestry subset.** Burden of rare TREs overlapping fetal heart epigenetic marks in individuals with TOF compared with controls in the European-ancestry subset. Bars represent odds ratios from Fisher's exact tests; error bars indicate 95% confidence intervals. The horizontal dashed line indicates an odds ratio of 1.

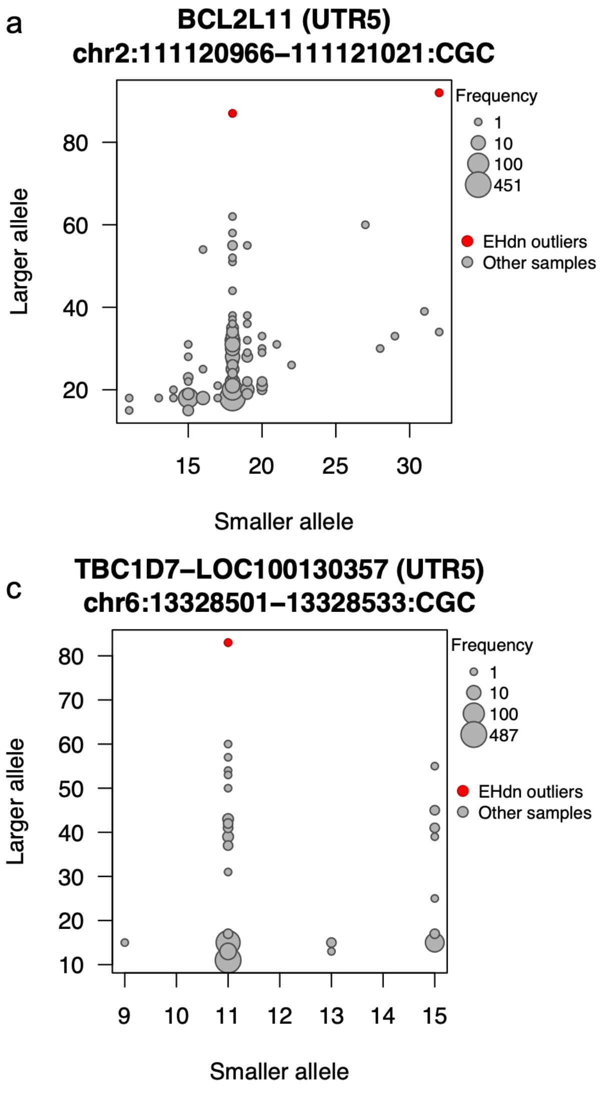


**Supplementary Figure 5. ExpansionHunter repeat-size distributions at prioritized 5′UTR tandem repeats.** Distribution of repeat sizes estimated using catalogue-based ExpansionHunter genotyping at four prioritized 5′UTR tandem repeat loci identified in the TOF cohort and presented in Table 1. (a–d) Repeat-size distributions at *BCL2L11* (a), *FAM149A* (b), *TBC1D7-LOC100130357* (c), and *ZNF713* (d). The x- and y-axes represent the repeat sizes of the two alleles estimated by ExpansionHunter. Each dot represents samples with the corresponding pair of allele repeat sizes, with dot size proportional to the number of samples. Samples identified as expansion outliers by ExpansionHunter Denovo (EHdn) are shown in red.

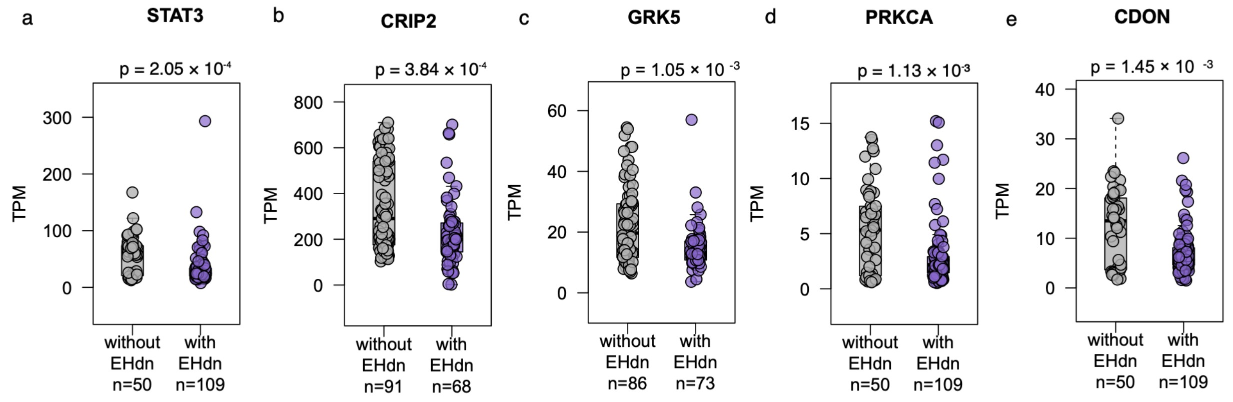


**Supplementary Figure 6. Gene expression differences associated with large tandem repeats in individuals with tetralogy of Fallot.** Expression of **(a)** *STAT3*, **(b)** *CRIP2*, **(c)** *GRK5*, **(d)** *PRKCA*, and **(e)** *CDON*, measured as transcripts per million (TPM), in individuals with and without a large repeat, as identified by EHdn. P-values are from two-sided Wilcoxon rank-sum tests. EHdn, ExpansionHunter Denovo.

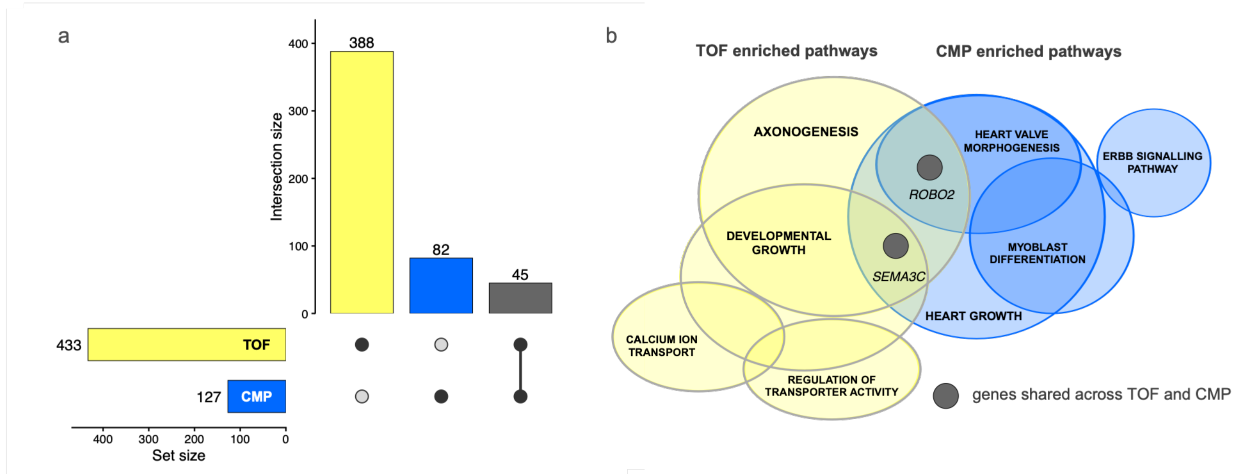


**Supplementary Figure 7. Comparison of tandem repeat expansion-associated genes and biological processes between tetralogy of Fallot and cardiomyopathy.**
**(a)** UpSet plot showing the overlap between genes associated with rare TREs in tetralogy of Fallot (TOF) and cardiomyopathy (CMP). **(b)** Enriched biological processes associated with TRE genes in TOF (yellow) and CMP (blue). Overlap within each cohort indicates shared genes. Circle sizes are adjusted to accommodate shared genes and do not represent a quantitative measure. Genes are shown in dark grey only when shared across the TOF and CMP cohorts.
